# Ultra-processed foods and type 2 diabetes mellitus incidence in RaNCD project: A prospective cohort study

**DOI:** 10.1101/2024.05.27.24307997

**Authors:** Parsa Amirian, Mahsa Zarpoosh, Yahya Pasdar

## Abstract

**Background:** Following rapid population growth and urbanization, global ultra-processed food consumption levels have increased. Additionally, type 2 diabetes mellitus, a non-communicable disease, is affecting one-tenth of the people worldwide. In this study, we aimed to investigate the association between ultra-processed food consumption and the risk of type 2 diabetes mellitus in different scenarios in a prospective cohort study in the western part of Iran.

**Methods:** The RaNCD cohort includes 10047 participants aged 35 to 65; the main phase commenced in March 2015; we included participants susceptible to diabetes at enrolment with follow-up data. We used the widely accepted NOVA classification to define ultra-processed foods. A multivariable Cox proportional hazards regression model was used as the main model; furthermore, the Cox model with different adjustments and the logistic regression model were used as sensitive analysis to evaluate the association between ultra-processed foods consumption and type 2 diabetes mellitus.

**Results:** A total of 8827 participants with a mean age of 46.92y, a mean follow-up time of 7.1y, and a mean daily ultra-processed food intake of 87.69 grams were included. During the follow-up phases, we included 255 incidences of type 2 diabetes mellitus cases. After adjusting for cofounders in the primary model, including age, gender, residence type, socioeconomic status, physical activity, body mass index, and familial history of diabetes despite the elevated hazard ratio of 1.08 (0.75, 1.55) in the fourth quartile compared to the first quartile, the P-value was insignificant (p-value = 0.665); p for trend in the UPF quartiles was also insignificant.

**Conclusion:** Our study has shed light on the association between ultra-processed food consumption and the risk of type 2 diabetes mellitus in the Middle East region. We have identified significant associations between diabetes incidence and some covariates. However, further investigations are necessary to confirm or refute the UPFs/T2DM association.

## Introduction

Since the advent of the NOVA classification and subsequently categorizing foods into four groups based on level of processing, including unprocessed, processed culinary ingredients, processed, and highly processed foods (ultra-processed foods (UPFs)), the effects of these ultra-processed foods on health, that are basically made of food constituents and food extracted substances enriched with flavors, enhancers, colors, and additives has become a major concern globally (1,2). The UPFs group has extremely diverse subgroups ranging from ultra-processed bread, flavored milk, margarine, and hydrogenated oils to pizzas, hamburgers, and ready-to-eat meals (3).

Due to rapid population growth and urbanization worldwide, industrial large-scale food production has become the primary way of producing foods in many countries; these large-scale food factories almost always produce UPFs; as the result of growing trends in UPFs consumption, scrutinizing the effects of UPFs consumption on health is essential (4-6). By affecting almost 10.5% of the world population, type 2 diabetes mellitus (T2DM) is a significant public health challenge, especially in regions with the fastest acceleration in T2DM prevalence, like the Middle East and North Africa (7,8). The association between UPFs and non-communicable diseases (NCDs), namely cardiovascular diseases (CVDs), obesity, hypertension, and dyslipidemia, has been shown previously (2,9-11). The association between UPFs and T2DM as an NCD has been investigated in countries worldwide, including Spain, the Netherlands, France, the United Kingdom, the United States, and South Korea. Still, these are mostly Western and developed countries, and none are located in the Middle East or North Africa, which, as previously said, have the fastest growth in T2DM prevalence (12-16). Because of the diverse methods of producing UPFs, different levels of processing, alterations in diets, and racial differences in each country, it is crucial to assess UPFs and NCDs separately in various parts of the world (17). To the best of our knowledge, our cohort study is the first in the Middle East region to assess the association between UPF consumption and T2DM incidence. The Ravansar non-communicable disease (RaNCD) cohort is part of the PERSIAN mega cohort, considered one of the biggest mega cohorts in the Middle East; RaNCD is the first cohort to assess the Kurdish population in western Iran (18). RaNCD uses a 113-item Food Frequency Questionnaire (FFQ) for nutritional status, and overall, 1186 variables in different domains for each participant. RaNCD’s main phase commenced in March 2015, and till May 2023, six successful follow-up phases have been completed (18). This study was conducted to fill the lack of knowledge regarding T2DM predisposing factors, protective factors, T2DM incidence, and an overall view of the UPF consumption trends in the Middle East region.

## Methods

### Study population

Using RaNCD cohort data, we performed a prospective analysis on 10047 participants aged between 35-65 years who reside in Ravansar County in the northwest of Kermanshah province in urban and rural areas (18). RaNCD cohort mainly focuses on NCDs, including T2DM, hypertension (HTN), CVDs, metabolic syndrome, …; the data collected by different means, namely laboratory tests, anthropometry, physical examination, and questionnaires, the collected bio-samples consists of blood, urine, hair, and nails; all procedures were performed in line with international guidelines (18). Initially, we reviewed data of 10,047 RaNCD cohort participants; we excluded participants with the missing data in the initial main phase (N = 232) in different variables, including fasting blood sugar (FBS), body mass index (BMI), socioeconomic status (SES), … two participants had missing data in the T2DM follow-up. We also excluded participants with improbable calorie intake <800 (N=32) or >5500 (N = 100). We also excluded participants who were diagnosed with T2DM already, were prescribed insulin, or were using diabetes-related drugs (N = 854). We found no pattern for our missing data; they were almost always completely random. Finally, we conducted our analysis of the data of 8827 participants (Figure 1 file S1).

**Figure 1.**
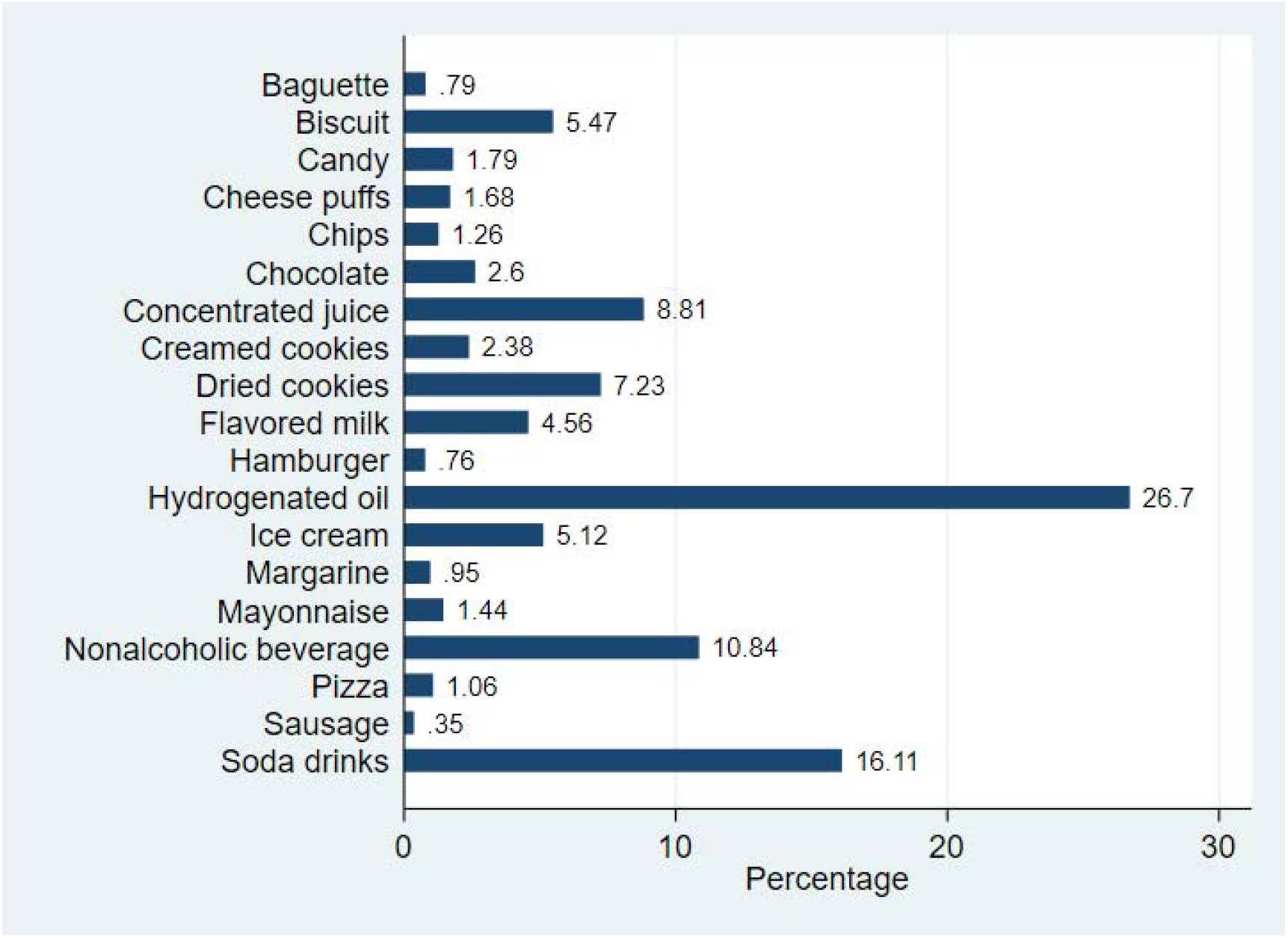
Each UPF percentage of contribution to the total UPF intake.

### Nutritional status & Ultra-processed foods

RaNCD uses the national Iranian Food Frequency Questionnaire (FFQ) to investigate the nutritional facts. We were provided with 113 baseline variables gathered using PERSIAN cohort FFQ and four local food baseline variables; these variables were reported by gram/day unit. We classified these 117 variables into NOVA groups (unprocessed, processed culinary ingredients, processed, and UPFs) (Table 3 file S1); the results were as follows: 65 variables were classified as unprocessed, 11 as processed culinary ingredients, 22 as processed, and 19 as UPFs; 4 local food variables wheat, cooked traditional greens, traditional bread, and Kermanshahi ghee were classified as 1, 2, 3, and 3 NOVA groups respectively. The identified UPF groups were as follows: 1. Baguette bread 2. Sausage/ salami 3. Hamburger 4. Pizza 5. Flavored milk 6. Margarine 7. Hydrogenated oil 8. Mayonnaise 9. Rock candy/ other sweets 10. Soda drinks 11. Nonalcoholic malt beverages 12. Ice cream 13. Dried cookies 14. Creamed cookies 15. Chocolate 16. Chips 17. Cheese puffs 18. Concentrated juice 19. Crackers/ biscuits. All UPF groups’ intakes were calculated previously as g/d units (Figure 1).

### Type 2 diabetes

During the six follow-up phases, we identified 267 participants who reported that they were diagnosed with T2DM; we excluded 12 participants from the analysis due to missing data; finally, we included 255 T2DM-positive participants in the statistical analysis; furthermore, calculated T2DM time at risk for all participants was 22,902,004 days. After participants reported they were diagnosed with T2DM by healthcare professionals, further assessment, including checking for fasting blood sugar (FBS), was conducted; participants with FBS >= 126 mg/dl during the follow-up period were considered new cases of T2DM. Figure 2 shows the cumulative hazard estimate of T2DM over time across UPF quartiles.

**Figure 2.**
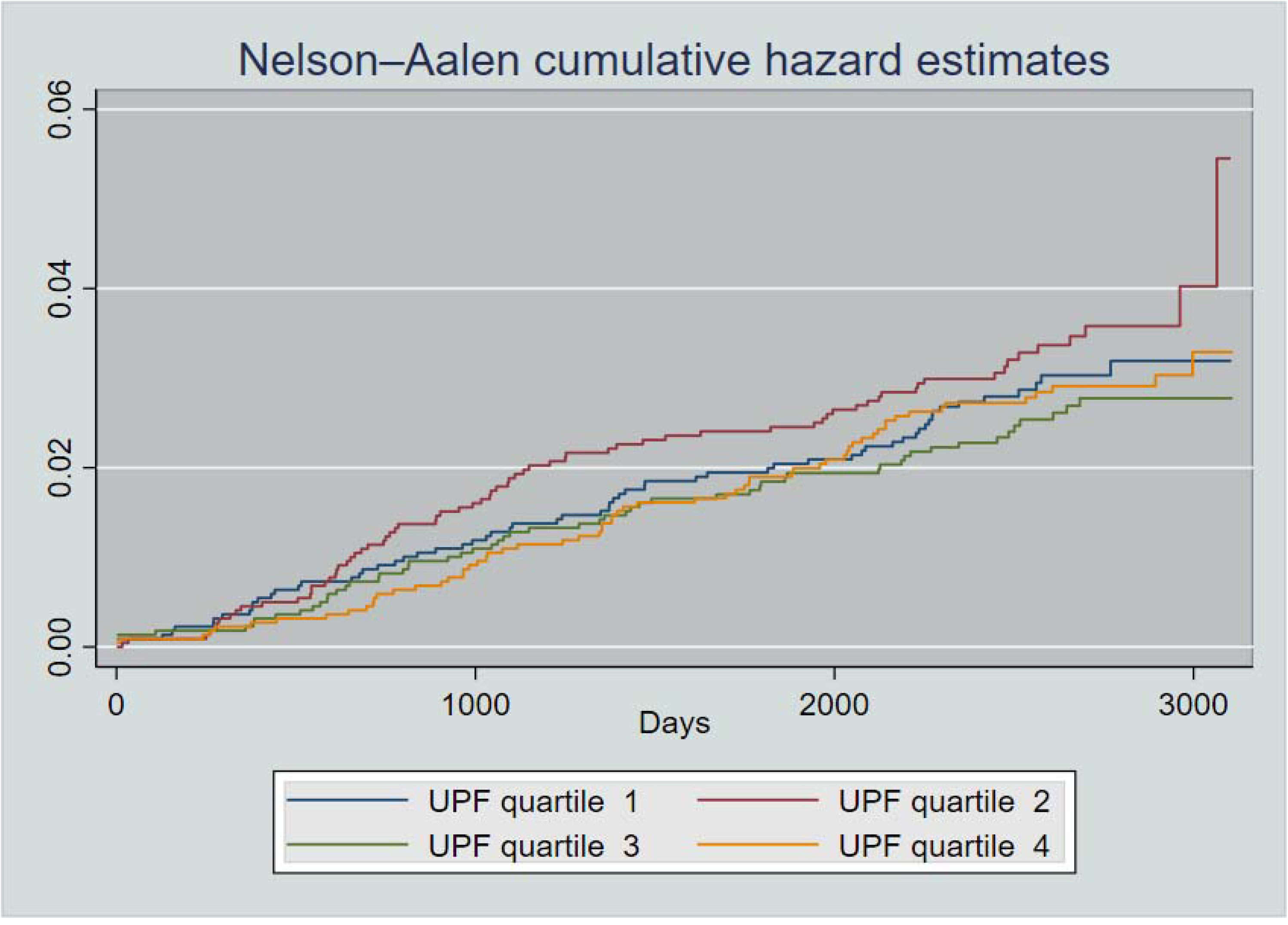
Nelson-Aalen curve showing hazard estimates of UPF quartiles.

### Statistical Analysis

We gathered data on different covariates for our main model and sensitivity analysis. Here is the list of our covariates: gender (male, female), age, residence type (urban or rural), marital status (single, not single), BMI, socioeconomic status (SES) quantiles, HTN history, CVD history, metabolic equivalent (MET) (low, moderate, high), familial history of T2DM, total lipid/ fat intake (g/d), total protein intake (g/d), total carbohydrate intake (g/d), total energy (kcal), alcohol use, smoking status (no, current, former, and passive), history of gastroesophageal reflux disease (GERD), history of blood in stool, history of muscle weakness, history of osteoporosis, history of rheumatoid arthritis, familial history of HTN, familial history of cardiac diseases, familial history of myocardial infarction (MI), and familial history of stroke. By adding values of every identified UPF item, we estimated each participant’s total grams of UPF consumption in a single day. Additionally, we stratified participants into quartiles based on their total grams of UFPs consumption (group one had the least consumption, and group four had the highest amount). Multivariable Cox proportional hazard models were used to analyze data to estimate hazard ratios (HRs) and 95% confidence intervals (CIs) in the main model and sensitivity analysis. In the main model, potential confounders include gender, age, residence type, marital status, BMI, SES quantiles, HTN history, MET, and familial history of T2DM. Several tests, including the Grambsch-Therneau test of proportional hazards, the calculation of concordance statistics, and the bootstrapping procedure, have been conducted to validate our results and ensure the robustness of our models. Furthermore, we designed nine different scenarios to assess the sensitivity and validity of our model. In scenario A, we used a logistic regression model to navigate our model under different modeling scenarios and assumptions; in scenario B, we adjusted main model for protein intake, lipid/fat intake, carbohydrate intake, and total energy; in scenario C we excluded early cases of T2DM incidence (<3 years), in scenario D we adjusted main model additionally for individual UPF items, in scenario E, we additionally adjusted main model for alcohol use. We stratified it for smoking status due to violating the model assumption. In scenario F, we investigated the effect of participants’ past medical histories (PMH)/symptoms on the main model. We inspected PMHs/symptoms: GERD, blood in stool, muscle weakness, osteoporosis, and rheumatoid arthritis (RA). In scenario G, we tuned the main model to assess the effects of familial histories, namely HTN, cardiac diseases, MI, and stroke. In scenario H, we checked for interactions between variables; six different interactions were evaluated (UPF quartile & age, gender & BMI, UPF quartile & gender, HTN & CVD, gender & MET groups, and UPF quartile & SES quantiles). In the last scenario (scenario I), we performed subgroup analysis; four subgroups were created (male or female, city or rural residence type, positive or negative history of CVD, and positive or negative familial history of T2DM).

## Results

### Participant characteristics & Main model

In this study, 8827 participants were included (47.39% male, 52.61% female), the mean age ± SD was 46.92y ± 8.23, the mean time of follow-up ± SD was 7.10y ± 1.24, 58.77% of participants were resided in urban areas (Table 1). The mean ± SD daily UPF intake among participants was 87.69 gr ± 84.24; participants with higher UPF consumption were more likely to be men, younger, reside in urban areas, were in upper SES quantiles, were more likely to be former/passive/current smoker, consume more energy, were less likely to have a positive history of CVD & HTN, had more physical activity, and had almost the same BMI & waist to hip ratio than the first UPF quartile.

**Table 1.** Characteristics of participants according to UPF quartiles (N = 8827)

| Characteristics | Quartile 1 (N = 2207) | Quartile 2 (N = 2207) | Quartile 3 (N = 2207) | Quartile 4 (N = 2206) |
| --- | --- | --- | --- | --- |
| | Mean ± SD or N (percentage) | | | |
| UPFs intake (g/d) | 19.62 ± 9.43 | 47.25 ± 7.81 | 83.46 ± 14.56 | 200.49 ± 95.01 |
| Age (years) | 48.86 ± 8.42 | 47.60 ± 8.22 | 46.13 ± 7.95 | 45.09 ± 7.80 |
| Male | 785 (35.57) | 898 (40.69) | 1129 (51.16) | 1371 (62.15) |
| Marital status |  |  |  |  |
| Single | 265 (12.01) | 261 (11.83) | 182 (8.25) | 155 (7.03) |
| Residence type |  |  |  |  |
| Urban | 1133 (51.34) | 1097 (49.71) | 1340 (60.72) | 1618 (73.35) |
| Alcohol use |  |  |  |  |
| No | 2173 (98.46) | 2141 (97.01) | 2091(94.74) | 2002 (90.75) |
| BMI | 27.59 ± 4.53 | 27.21 ± 4.56 | 27.23 ± 4.43 | 27.41 ± 4.52 |
| Smoking |  |  |  |  |
| No | 1003 (45.45) | 1003 (45.45) | 880 (39.87) | 769 (34.86) |
| Current | 177 (8.02) | 210 (9.52) | 277 (12.55) | 364 (16.50) |
| Former | 168 (7.61) | 162 (7.34) | 206 (9.33) | 187 (8.48) |
| Passive | 859 (38.92) | 832 (37.70) | 844 (38.24) | 886 (40.16) |
| SES quantiles |  |  |  |  |
| 1 | 540 (24.47) | 563 (25.51) | 399 (18.08) | 271 (12.28) |
| 2 | 473 (21.43) | 449 (20.34) | 441 (19.98) | 396 (17.95) |
| 3 | 432 (19.57) | 411 (18.62) | 435 (19.71) | 499 (22.62) |
| 4 | 368 (16.67) | 389 (17.63) | 463 (20.98) | 518 (23.48) |
| 5 | 394 (17.85) | 395 (17.90) | 469 (21.25) | 522 (23.66) |
| HTN |  |  |  |  |
| Yes | 400 (18.12) | 312 (14.14) | 275 (12.46) | 257 (11.65) |
| CVDs |  |  |  |  |
| YES | 440 (19.94) | 325 (14.73) | 281 (12.73) | 239 (10.83) |
| MET (group) |  |  |  |  |
| Low | 663 (30.04) | 664 (29.18) | 654 (29.63) | 669 (30.33) |
| Moderate | 1152 (52.20) | 1097 (49.71) | 1004 (45.49) | 963 (43.65) |
| High | 392 (17.76) | 466 (21.11) | 549 (24.88) | 574 (26.02) |
| Diabetes (FH) |  |  |  |  |
| Yes | 524 (23.74) | 517 (23.43) | 528 (23.92) | 498 (22.57) |
| Energy (kcal) | 2168.70 ± 734.99 | 2430.27 ± 736.29 | 2794.15 ± 765.78 | 3326.86 ± 882.58 |

The test of the proportional hazards assumption showed no violation. Our main model’s C-index was 0.7517, and the bootstrap results were identical to the main model.

The hazard ratio (95% CI) of T2DM in the highest UPF quartile was 1.08 (0.75, 1.55) compared to the first quartile, but the P-value was insignificant (p-value = 0.665). However, our Cox proportional hazard model revealed several statistically significant associations. The hazard ratio (95% CI, p-value) of age on T2DM was 1.03 (1.02, 1.05, p-value < 0.001), the hazard ratio (95% CI, p-value) of living in rural areas on T2DM was 0.66 (0.48, 0.90, p-value = 0.008), the hazard ratio (95% CI, p-value) of BMI on T2DM was 1.14 (1.11, 1.16, p-value < 0.001), the hazard ratio (95% CI, p-value) of positive history CVD on T2DM was 1.54 (1.03, 2.29, p-value = 0.032), the hazard ratio (95% CI, p-value) of positive familial history of T2DM was 1.84 (1.39, 2.44, p-value <0.001); other covariates were not statistically associated with T2DM.

### Sensitivity analysis

To test our hypothesis under different assumptions, we conducted our analysis using the Logistic regression model; Table 2 show the HRs and 95% CIs of age, BMI, familial history of diabetes, and each UPF quartile under different models. Our adjustment for total daily protein intake, total carbohydrate, total lipid/fat, and total energy showed no significance; by excluding early cases (<3y), we found that the hazard ratio of positive history of HTN was 1.66 (1.00, 2.77, p-value = 0.049). Still, the results of a positive history of CVD became insignificant (p-value = 0.427). We fit our main model for each UPF item; the only UPF item with a significant hazard ratio (95%CI, p-value) was mayonnaise 1.04 (1.00, 1.09, p-value = 0.046). We also explored the effects of alcohol use and smoking status on T2DM incidence; the smoking status variable did not hold the Cox model assumption, and because of that, we stratify the main model to smoking status and further adjust the alcohol consumption as a cofounder by doing so the results did not change meaningfully. We also checked for the hazard ratios related to PMHs; surprisingly, we found that muscle weakness had a hazard ratio (95%CI, p-value) of 0.18 (0.04, 0.74, p-value = 0.018). Subsequently, we checked for the familial history of HTN, cardiac diseases, MI, and stroke; we found that participants with a positive history of stroke in first-degree relatives had a hazard ratio (95%CI, p-value) of 1.51 (1.11, 2.05, p-value = 0.007). Assessing the interactions between variables provided in the scenario, H showed no significance except for the interaction between gender and BMI; the hazard ratio (95%CI, p-value) for the mentioned interaction was 0.94 (0.88, 0.99, p-value = 0.029) (Table 1 file S1). Lastly, we performed subgroup analysis (Table 2 file S1); the comparison between males and females showed that the hazard ratio of residence type is not significant, and the hazard ratio (95%CI, p-value) of HTN was 1.60 (1.00, 2.55, p-value = 0.049). Still, the hazard ratio of CVD became statistically insignificant. The comparison between participants living in urban and rural areas showed that participants living in cities and were in third-level SES had a hazard ratio (95%CI, p-value) of 0.57 (0.34, 0.95, p-value = 0.033); interestingly, participants who were living in rural areas and were in second MET group compared to first MET group had hazard ratio (95%CI, p-value) of 0.49 (0.28, 0.85, p-value = 0.012) for T2DM incidence. Participants with negative history of CVD in the second UPF quartile had a hazard ratio (95%CI, p-value) of 1.58 (1.04, 2.41. p-value = 0.030) compared to the first quartile; additionally, participants with negative familial history of T2DM, in the second UPF quartile had a hazard ratio (95%CI, p-value) of 1.60 (1.03, 2.48. p-value = 0.036) compared to the first quartile. Participants who were not single (married/divorced/widowed) with a positive familial history of T2DM had a hazard ratio (95%CI, p-value) of 3.62 (1.12, 11.6, p-value = 0.031).

**Table 2.**
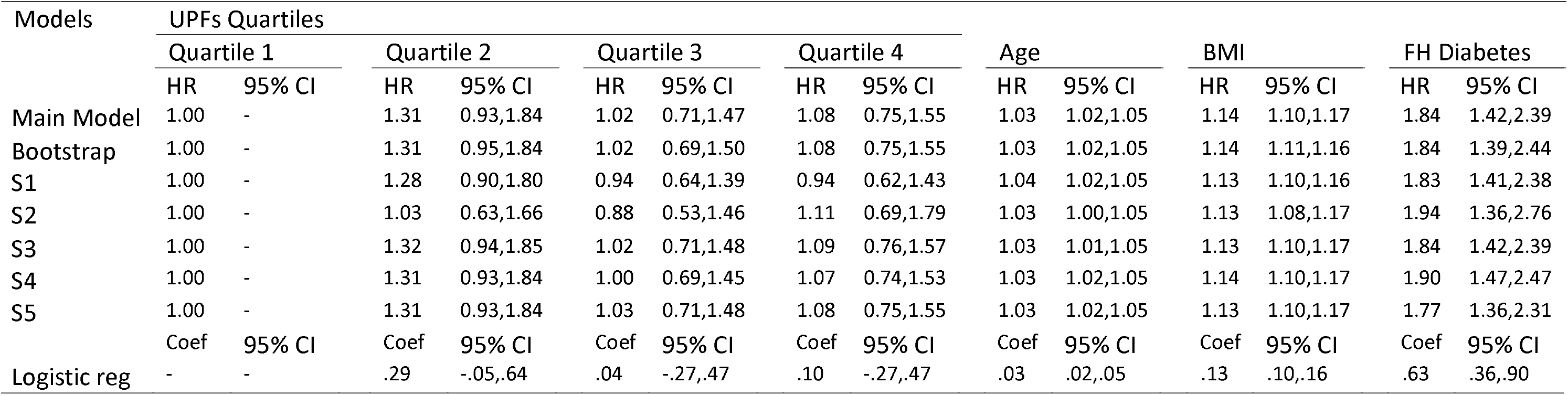
HRs and 95% CIs of age, BMI, familial history of diabetes, and each UPF quartile under different models. All p-values associated with UPFs quartiles were insignificant (p-value > 0.05), adversely all p-values associated with age, bmi, and FH diabetes were significant (p-value < 0.05). S1: Main model + total protein + total lipid + total carbohydrate + Energy S2: Excluding early cases (< 3y) S3: Main model + alcohol use & stratification by smoking status S4: Main model + GERD history + blood in stool history + muscle weakness history + osteoporosis history + rheumatoid arthritis history S5: Main model+familial histories (HTN, CVDs, Ml, stroke)

## Discussion

In the RaNCD prospective cohort of Iranian/Kurdish adults aged between 35 and 65y, we found no significant association between UPF consumption and T2DM. Even after creating quartiles of UPF consumption, we detected no significance in both cases; HRs were >1, but p-values did not show significance; our test to calculate p for trend using medians also showed no associations. Generally speaking, developing countries consume fewer UPFs (19); this is particularly the case in rural areas and small cities in these countries like Ravansar County; we think that is why the mean percentage of UPF intake in grams was only 3.0% of total grams of foods and beverages consumed. Compared to other countries namely South Korea (4.9%) (16), Spain (9.5%) (14), France (17.3%) (15), United Kingdom (22.1%) (13), and United States (36.1%) (12) it is the lowest. The results lead us to assume that consuming UPFs in these quantities (3% of total food intake) is not associated with T2DM incidence. Another explanation for not seeing an association might be because we strictly tried to detect and adjust potential confounders like BMI; we aimed to assess the direct effect of UPF consumption, not by indirect paths (e.g., increasing BMI); after adjusting for BMI, Canhada et al. also found no significant association (20). We also adjusted for familial history of T2DM in first-degree relatives to control for genetic susceptibilities, which is shown to affect T2DM incidence greatly (21). NOVA food classification is a qualitative classification; the level of processing pizza as a UPF, for example, is undoubtedly different among countries, yet we all assume it as UPF; our UPFs in Iran may be less processed than our counterparts, and this might be a part of the reason that we found no association, unlike other studies. The weight ratio of unprocessed foods was more than half of total food intake (59.51%), Processed Culinary Ingredients were responsible for 10.25% of total food intake, the weight ratio of processed foods was 27.16%, and lastly UPFs proportion was only 3.08% (Figure 3); part of the reason that UPFs weight ratio is minimal in our study might be because we tried to be extremely cautious on entering different foods to UPFs group that is why we did not consider canned tuna, molasses, yogurt, and coffee as UPFs.

**Figure 3.**
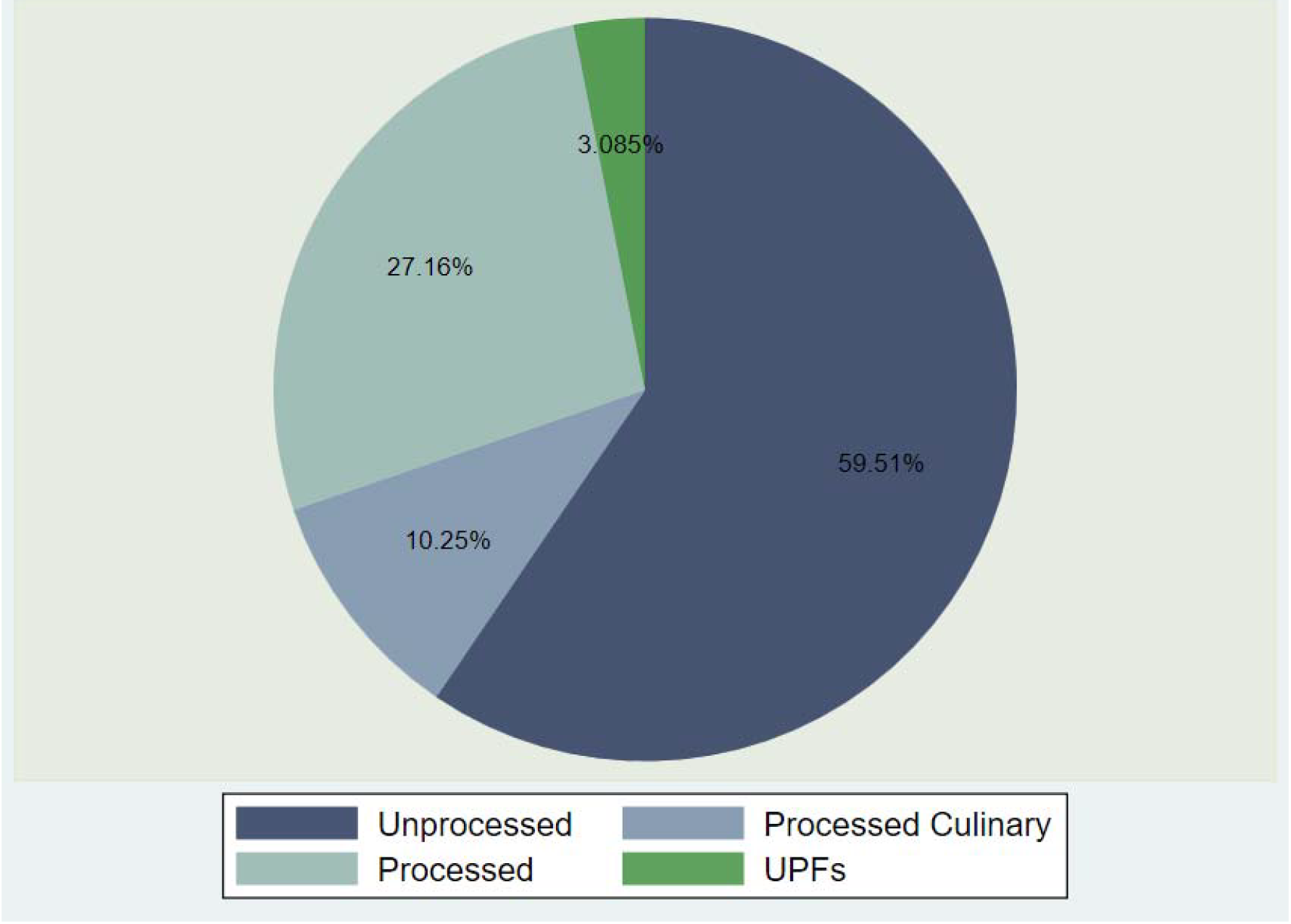
Contribution of each NOVA group to mean weight of foods consumed.

There is still a massive gap in our understanding of whether UPF consumption can lead to an increased risk of T2DM. UPFs are enriched with emulsifiers, nonnutritive sweeteners, and enhancers; studies that directly assess the role of these additives can be crucial to improving our understanding of underlying mechanisms; one study showed that mice that were treated with carrageenan food additive had substantially impaired glucose tolerance and were more resistant to insulin (22). Other than the UPFs/T2DM relation, our models showed some significant associations. We found that if other factors remain constant for each additional year of age, the risk of T2DM incidence increases by 3.83%; we also found that participants who reside in rural areas of Ravansar county had a 33.68% decreased chance of T2DM than their counterparts who reside in urban areas; BMI also was a substantial predictor, our model showed that for 1 unit increase in BMI, there is a 14.04% increased chance of T2DM incidence, interestingly we found that positive history of CVD increases the chance of T2DM incidence by 54.47%; lastly we found in Iranian/Kurdish population with positive familial history of T2DM and stroke in first-degree relatives the chance of T2DM increases by 84.91% and 51.70% respectively. In the sensitivity analysis, when excluding the early cases (<3y), we found that HTN history increases the chance of T2DM incidence by 66.91%, but CVDs statistically do not increase the chance (p-value = 0.427). Participants who reported muscle weakness in a part of the body lasting more than a week statistically had an 81.65% decreased chance of T2DM. By checking the interaction between gender and BMI, we found that BMI as a risk factor operates slightly differently in different genders, suggesting that females have slightly decreased risk (6%). In the subgroup analysis, we found that in men, there is a significant association between CVDs and T2DM with a more than twofold increase in hazard (154%). We also found that women with a positive history of HTN are 60% more at risk.

We have faced several limitations in conducting the current study; first, the PERSIAN FFQ is not specifically designed to distinguish UPFs from other NOVA groups; we manually categorized 117 food items of PERSIAN FFQ into NOVA groups; second, our follow-up time (8.5 years) is relatively a short period for life-long chronic diseases like T2DM to unravel, the results may change as the follow-up period expands. Third, although we strictly tried to identify potential confounders, there is still a chance that unmeasured confounders/residual confounding alters the results. Fourth, the incidence of T2DM was documented by participants’ self-report; this can introduce a potential bias to the analysis.

Our study stands as a pioneering effort in the Middle East and North Africa region and, more broadly, among developing countries. We meticulously designed nine different scenarios in the sensitivity analysis, ranging from using various statistical models to checking for interactions and subgroup analysis. Beyond the UPFs/T2DM association, we unearthed significant HRs for T2DM incidence in our population. Lastly, our cohort participants represent the actual demographic of the population, with a diverse mix of occupations, SESs, and residence locations, adding to the richness of our findings.

In conclusion, our study sheds light on the association of UPFs/T2DM in the Middle East. We have also identified significant associations between T2DM incidence and some covariates. However, it is crucial to note that our findings underscore the need for further investigations. The UPFs/T2DM association is a complex issue that requires more comprehensive studies to be definitively proven or disproven.

## Data Availability

All data produced in the present study are available upon reasonable request to the authors

## Ethics Approval

Ethical approval for the study was obtained from the Kermanshah University of Medical Sciences Ethics Committee.

## Acknowledgments

We thank the participants of the RaNCD cohort for their cooperation and participation. We also want to thank all members of the RaNCD cohort for their unwavering support.

