## Supplementary files for "Ultra-processed foods and type 2 diabetes mellitus incidence in RaNCD project: A prospective cohort study"

| Interactions |  | | | | | | | | | | |  |  | |  |  | |
| --- | --- | --- | --- | --- | --- | --- | --- | --- | --- | --- | --- | --- | --- | --- | --- | --- | --- |
|  | UPFq & Age | |  | Gender & BMI | |  | UPFq & Gender | |  | HTN & CVD | |  | Gender & METg | |  | UPFq & SESq | |
|  | HR | 95% CI |  | HR | 95% CI |  | HR | 95% CI |  | HR | 95% CI |  | HR | 95% CI |  | HR | 95% CI |
| UPFq1 & Age | _ | _ |  |  |  |  |  |  |  |  |  |  |  |  |  |  |  |
| UPFq2 & Age | 1.02 | 0.98,1.06 |  |  |  |  |  |  |  |  |  |  |  |  |  |  |  |
| UPFq3 & Age | 1.03 | 0.98,1.07 |  |  |  |  |  |  |  |  |  |  |  |  |  |  |  |
| UPFq4 & Age | 1.01 | 0.97,1.05 |  |  |  |  |  |  |  |  |  |  |  |  |  |  |  |
| Gender1 & BMI |  |  |  | _ | _ |  |  |  |  |  |  |  |  |  |  |  |  |
| Gender2 & BMI |  |  |  | 0.94 | 0.88,0.99 |  |  |  |  |  |  |  |  |  |  |  |  |
| UPFq1 & Gender1 |  |  |  |  |  |  | _ | _ |  |  |  |  |  |  |  |  |  |
| UPFq2 & Gender2 |  |  |  |  |  |  | 1.30 | 0.64,2.63 |  |  |  |  |  |  |  |  |  |
| UPFq3 & Gender2 |  |  |  |  |  |  | 1.71 | 0.81,3.60 |  |  |  |  |  |  |  |  |  |
| UPFq4 & Gender2 |  |  |  |  |  |  | 1.33 | 0.65,2.73 |  |  |  |  |  |  |  |  |  |
| HTN & CVD |  |  |  |  |  |  |  |  |  | 0.91 | 0.44,1.88 |  |  |  |  |  |  |
| Gender1 & METg1 |  |  |  |  |  |  |  |  |  |  |  |  | _ | _ |  |  |  |
| Gender2 & METg2 |  |  |  |  |  |  |  |  |  |  |  |  | 1.82 | 0.97,3.40 |  |  |  |
| Gender2 & METg3 |  |  |  |  |  |  |  |  |  |  |  |  | 0.66 | 0.27,1.61 |  |  |  |
| UPFq1 & SESq1 |  |  |  |  |  |  |  |  |  |  |  |  |  |  |  | _ | _ |
| UPFq2 & SESq2 |  |  |  |  |  |  |  |  |  |  |  |  |  |  |  | 1.07 | 0.34,3.36 |
| UPFq2 & SESq3 |  |  |  |  |  |  |  |  |  |  |  |  |  |  |  | 1.10 | 0.38,3.14 |
| UPFq2 & SESq4 |  |  |  |  |  |  |  |  |  |  |  |  |  |  |  | 0.73 | 0.25,2.15 |
| UPFq2 & SESq5 |  |  |  |  |  |  |  |  |  |  |  |  |  |  |  | 0.76 | 0.25,2.22 |
| UPFq3 & SESq2 |  |  |  |  |  |  |  |  |  |  |  |  |  |  |  | 1.46 | 0.46,4.63 |
| UPFq3 & SESq3 |  |  |  |  |  |  |  |  |  |  |  |  |  |  |  | 0.46 | 0.13,1.54 |
| UPFq3 & SESq4 |  |  |  |  |  |  |  |  |  |  |  |  |  |  |  | 0.55 | 0.17,1.74 |
| UPFq3 & SESq5 |  |  |  |  |  |  |  |  |  |  |  |  |  |  |  | 0.48 | 0.15,1.56 |
| UPFq4 & SESq2 |  |  |  |  |  |  |  |  |  |  |  |  |  |  |  | 0.94 | 0.28,3.13 |
| UPFq4 & SESq3 |  |  |  |  |  |  |  |  |  |  |  |  |  |  |  | 0.42 | 0.13,1.39 |
| UPFq4 & SESq4 |  |  |  |  |  |  |  |  |  |  |  |  |  |  |  | 0.64 | 0.21,1.95 |
| UPFq4 & SESq5 |  |  |  |  |  |  |  |  |  |  |  |  |  |  |  | 0.50 | 0.16,1.60 |

Table 1 HRs and CIs associated with each interaction; all p-values except for BMI & gender were insignificant (p-value > 0.05).

Table 2 HRs and 95% CIs of age, BMI, familial history of diabetes, and each UPF quartile in the subgroup analysis.

* All p-values associated with UPFs quartiles were insignificant ( p-value > 0.05) except for starred ones; p-values associated with age, BMI, and familial history of diabetes were significant except for starred ones ( p-value < 0.05).

| Subgroup Analysis | UPFs Quartiles | | | | | | | | | | |  |  | |  |  | |  |  | |
| --- | --- | --- | --- | --- | --- | --- | --- | --- | --- | --- | --- | --- | --- | --- | --- | --- | --- | --- | --- | --- |
|  | Quartile 1 | |  | Quartile 2 | |  | Quartile 3 | |  | Quartile 4 | |  | Age | |  | BMI | |  | FH Diabetes | |
|  | HR | 95% CI |  | HR | 95% CI |  | HR | 95% CI |  | HR | 95% CI |  | HR | 95% CI |  | HR | 95% CI |  | HR | 95% CI |
| Gender |  |  |  |  |  |  |  |  |  |  |  |  |  |  |  |  |  |  |  |  |
| Male | 1.00 | _ |  | 1.09 | 0.62,1.92 |  | 0.70 | 0.38,1.27 |  | 0.89 | 0.51,1.56 |  | 1.04 | 1.01,1.07 |  | 1.18 | 1.13,1.24 |  | 2.07 | 1.36,3.16 |
| Female | 1.00 | _ |  | 1.41 | 0.92,2.16 |  | 1.23 | 0.77,1.96 |  | 1.18 | 0.73,1.91 |  | 1.03 | 1.01,1.05 |  | 1.11 | 1.07,1.15 |  | 1.76 | 1.27,2.46 |
| Age |  |  |  |  |  |  |  |  |  |  |  |  |  |  |  |  |  |  |  |  |
| >50 | 1.00 | _ |  | 1.87* | 1.15,3.03 |  | 1.41 | 0.82,2.45 |  | 1.17 | 0.65,2.10 |  |  |  |  | 1.12 | 1.07,1.16 |  | 1.82 | 1.21,2.73 |
| <50 | 1.00 | _ |  | 0.86 | 0.53,1.39 |  | 0.68 | 0.42,1.12 |  | 0.85 | 0.53,1.35 |  |  |  |  | 1.14 | 1.10,1.19 |  | 1.83 | 1.30,2.56 |
| Residence |  |  |  |  |  |  |  |  |  |  |  |  |  |  |  |  |  |  |  |  |
| Urban | 1.00 | _ |  | 1.16 | 0.77,1.75 |  | 0.80 | 0.51,1.24 |  | 0.98 | 0.65,1.46 |  | 1.03 | 1.01,1.05 |  | 1.13 | 1.09,1.17 |  | 1.98 | 1.47,2.68 |
| Rural | 1.00 | _ |  | 1.69 | 0.91,3.13 |  | 1.61 | 0.83,3.16 |  | 1.17 | 0.51,2.70 |  | 1.04 | 1.01,1.07 |  | 1.14 | 1.08,1.20 |  | 1.48* | 0.87,2.51 |
| CVD |  |  |  |  |  |  |  |  |  |  |  |  |  |  |  |  |  |  |  |  |
| Positive | 1.00 | _ |  | 0.82 | 0.44,1.51 |  | 1.16 | 0.64,2.12 |  | 0.67 | 0.33,1.38 |  | 1.0* | 0.98,1.04 |  | 1.10 | 1.05,1.16 |  | 1.25* | 0.74,2.09 |
| Negative | 1.00 | _ |  | 1.58* | 1.04,2.41 |  | 0.96 | 0.60,1.53 |  | 1.30 | 0.84,2.02 |  | 1.04 | 1.02,1.06 |  | 1.15 | 1.11,1.19 |  | 2.11 | 1.56,2.87 |
| FH diabetes |  |  |  |  |  |  |  |  |  |  |  |  |  |  |  |  |  |  |  |  |
| Positive | 1.00 | _ |  | 0.94 | 0.54,1.62 |  | 0.76 | 0.43,1.35 |  | 0.60 | 0.32,1.11 |  | 1.03 | 1.00,1.06 |  | 1.10 | 1.05,1.16 |  |  |  |
| Negative | 1.00 | _ |  | 1.60 | 1.03,2.48 |  | 1.2* | 0.74,1.95 |  | 1.45 | 0.92,2.31 |  | 1.03 | 1.01,1.06 |  | 1.15 | 1.11,1.19 |  |  |  |

| GROUP 1 – Unprocessed or minimally processed foods | | | |
| --- | --- | --- | --- |
| Fruits  Vegetables  Nuts  Diaries  Cereals  Meats  Others | apricot, sweet cherry, sour cherry, cantaloupe melon, watermelon, nectarine, peach, plums, fresh berries, strawberries, pomegranate, banana, fresh figs, persimmon, grapes, dates, pears, olives, apple, kiwi, citrus fruits, dried fruits, raisin, honeydew  cabbage, tomato, potato, carrot, fresh leafy greens, garlic, zucchini, cucumber, onion, beets, green peppers, bell peppers, celery, eggplant, lettuce, green beans, green peas, soybean, boiled fava beans or lima beans  peanut, walnut, seeds, other nuts  milk  wheat, wheat oats barley, corn, boiled lentils, boiled chickpeas, boiled split peas  boiled or grilled red meat, a variety of meats and their products, boiled chicken, boiled sheep brain, boiled sheep tongue, chicken giblet, boiled or grilled fish  boiled egg, mushroom, tea | | |
| GROUP 2 – Processed culinary ingredients | | | |
| Cooked  Oils  Sugars  Diaries  Others | | cooked leafy greens, cooked rice, cooked traditional greens  olive oils, other plant oils  white sugar, sugar cube  butter  fruit juice, honey, salt | |
| GROUP 3 – Processed foods | | | |
| Bread  Diaries  Conserves  Pickled  Oils  Others | | sangak bread, lavash bread, barbary bread, barely bread, traditional bread  yoghurt, cheese, dough, kashk, clotted cream  tomato pastes, pomegranate paste, jam, tuna canned, compotes  pickles, pickled vegetables, torshi  Kermanshahi ghee,  tahini halva, NESCAFE coffee, cooked pasta noodles | |
| GROUP 4 – Ultra-processed foods | | | |
| Drinks  Fast foods  Oils  Snacks  Dairies  Others | | | soda drinks, non-alcoholic beverages malt  Pizza, sausages or bologna, hamburger  margarine vegetable butter, hydrogenated oil of animal fats  crackers or wafers biscuits, candies or sweets, dry cookies, creamed cookies, chocolate, chips, cheese puffs  ice cream, flavored milk  mayonnaise salad dressing, concentrate juice, baguette bread |

Table 3 NOVA classification of PERSIAN cohort FFQ items.

Total participants in the RaNCD Cohort

(n=10047)

Missing data

(n=232)

Total participants after the removal of observations

With missing data

(n=9815)

Positive history of diabetes mellitus or anti-diabetic drug consumption

(n=854)

Total participants after

the removal of observations with a history of diabetes mellitus or anti-diabetic drugs

(n=8961)

Missing data in

T2DM follow-up

(n=2)

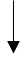

Total participants after the removal of missing data in T2DM follow-up

(n=8959)

Observations available for analyses in this study

(n=8827)

Implausible daily calorie intake

(n=132)

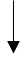

Figure 1 Flow-chart of participants.
